# Transient elastography assessed hepatic steatosis and fibrosis are associated with body composition in the United States

**DOI:** 10.1101/2020.05.01.20087510

**Authors:** Aynur Unalp-Arida, Constance E. Ruhl

## Abstract

**Background & Aims:** We examined transient elastography assessed hepatic steatosis and fibrosis distributions and relationships with body composition in a representative United States population sample.

**Methods:** Liver stiffness and controlled attenuation parameter (CAP) were assessed on 4,870 non-Hispanic white, non-Hispanic black, non-Hispanic Asian, and Hispanic men and women aged 20 years and over in the National Health and Nutrition Examination Survey (NHANES) 2017-2018. Participants underwent anthropometry and dual-energy x-ray absorptiometry (DXA).

**Results:** Compared to women, men had higher mean CAP (274.2 dB/m vs. 254.4 dB/m) and liver stiffness (6.4 kPa vs. 5.5 kPa). CAP and liver stiffness increased with age and BMI. In multivariate-adjusted analysis, CAP in the upper quartile was associated with increased age, BMI, waist-to-hip ratio, ALT and C-reactive protein (p<0.001 for each). After adjustment, non-Hispanic blacks had lower CAP and non-Hispanic Asians had over twice the odds of higher CAP. In multivariate-adjusted analysis, liver stiffness in the upper quartile was associated with male sex, increased age, BMI, GGT, and CAP (p<0.001 for each), and hepatitis C virus positivity. Lower stiffness among Non-Hispanic Asians was not significant after adjustment for BMI. DXA trunk and extremity fat mass were positively related to both CAP and liver stiffness with adjustment for sex, race-ethnicity, and age (p<0.001 for each). Results were similar with CAP and liver stiffness as continuous characteristics.

**Conclusion:** In the U.S. population, increased anthropometric and DXA body composition measures were associated with higher CAP and liver stiffness. Racial-ethnic differences observed merit continuation of NHANES transient elastography to further elucidate the burden of obesity and liver health disparities.

## INTRODUCTION

The obesity epidemic continues to be a significant public health burden in the United States with doubling prevalence of severe obesity from 4.7% to 9.2% and steady increases in the age-adjusted prevalence of all obesity from 30.5% to 42.4% in the past two decades.^1^ Accompanying the obesity epidemic, chronic liver disease and cirrhosis are responsible for increased morbidity^2^ and ranked as the 11^th^ leading cause of death in the United States in 2017 (https://www.cdc.gov/nchs/data/nvsr/nvsr68/nvsr68_09-508.pdf). Noninvasive tests including transient elastography, body composition and blood biomarkers are practical tools to track and promote liver health in the general population.^3,4^

Current guidelines suggest noninvasive tests for the initial assessment of liver disease, specifically hepatic steatosis and fibrosis in primary health care settings.^5–7^ Among available noninvasive tests, transient elastography by FibroScan® (Echosens, Paris, France) is commonly used by health care practitioners worldwide for chronic liver disease surveillance over the past decade.^8–13^ Transient elastography measures liver stiffness and the controlled attenuation parameter (CAP) simultaneously and can serve as a one-stop examination for both liver steatosis and fibrosis intermediate outcomes.^14,15^ Although obesity is a common reason for transient elastography measurement failures, the development of the XL probe allows complete measures in the majority of obese patients.^16,17^ Transient elastography has a high diagnostic accuracy for identifying advanced fibrosis and cirrhosis and is also useful for measuring liver fat.^18^

Over the past two decades, fatty liver disease has been increasingly studied using data from the National Health and Nutrition Examination Survey (NHANES) due to its emergence as a growing public health burden. Analyses of various biomarkers for liver injury, including liver enzyme activities, abdominal ultrasound, and fat and fibrosis scores have estimated prevalence, identified associated factors, and demonstrated relationships with higher mortality.^19–22^ NHANES data have also been used to study body composition in the United States in relation to liver and other chronic diseases using dual-energy x-ray absorptiometry (DXA) measurements to develop anthropometric prediction equations for lean body mass, fat mass and percent fat in adults.^23,20^

Building on these epidemiological advances, transient elastography was used to measure hepatic steatosis and fibrosis on eligible participants in NHANES 2017-2018. These data are now available and provide the first U.S. nationally representative transient elastography assessed liver stiffness and CAP observations along with other available survey measures.^24,25^ We estimated the U.S. population distribution of hepatic steatosis and fibrosis biomarkers assessed by transient elastography and examined associated anthropometric and DXA body composition characteristics.

## MATERIALS AND METHODS

### Source population

The NHANES is conducted in the United States by the National Center for Health Statistics of the Centers for Disease Control and Prevention.^26^ The survey consists of cross-sectional interview, examination, and laboratory data collected from a complex multistage, stratified, clustered probability sample representative of the civilian, noninstitutionalized population with oversampling of non-Hispanic blacks, Hispanics, Asians, low income persons, and older adults. The survey was approved by the Centers for Disease Control and Prevention ethics review board, and all participants provided written informed consent. We utilized the NHANES 2017-2018 sample during which liver fat and stiffness biomarkers were assessed for the first time in NHANES by transient elastography.

### Transient elastography

Vibration controlled transient elastography using FibroScan® 502 V2 Touch (Echosens™ North America, Waltham, MA) was performed on eligible NHANES 2017-2018 participants according to the manufacturer guidelines.^24, 25, 27^ Participants 20 years and older were asked to fast for at least 3 hours prior to their mobile examination center visit. Multiple measurements up to ≥30 were made on each participant using a medium (M; 72% of participants) or large (XL) probe. Hepatic fibrosis was measured using transient elastography-derived liver stiffness in kilopascals (kPa) with median, interquartile range (IQR), and IQR/median calculated for each survey participant. Simultaneously, hepatic steatosis was measured using CAP in decibels per meter (dB/m) with median and IQR calculated for each participant. The inter-rater reliability (n=32) between NHANES health technicians and reference examiners (expert FibroScan^®^ technicians) was 0.86 for stiffness and 0.94 for CAP-steatosis with mean differences (standard deviation) for stiffness (kPa) of 0.44 (1.3), and for CAP-steatosis (dB/m) of 4.5 (19.8).

### Other variables

NHANES 2017-2018 data collected on demographic and clinical characteristics were examined in relation to liver stiffness and CAP. Age (years), sex, race-ethnicity (non-Hispanic white, non-Hispanic black, non-Hispanic Asian, Hispanic, or other), income, and liver condition history were ascertained by interview.^28,29^ Income was measured by the poverty income ratio (ratio of family income to poverty threshold) and categorized as <1.0, 1.0-<2.0, 2.0-<3.0, 3.0-<5.0 and ≥5.0. Poverty income ratio of 1.0 and 5.0 represent family incomes equal to 1 and 5 times, respectively, the poverty threshold for a given family size and year. Participants were asked if a doctor or other health professional ever told them that they had a liver condition, whether they still had a liver condition, and the age when first told they had a liver condition.

Body measurements including standing height (cm), weight (kg), waist circumference (cm), and hip circumference (cm) were ascertained during the mobile examination center visit, and body mass index (BMI; weight [kg]/height [m^2^]) and waist-to-hip circumference ratio were calculated.^30^ DXA body composition measures were available on participants aged 20-59 years.^31^ Arm, leg, and trunk fat mass (gm) and total percent fat (%) were measured by DXA by using Hologic Discovery model A densitometers (Hologic Inc., Bedford, MA); Hologic APEX version 4.0 software was used for analysis of the scans. Arm and leg fat mass summed to extremity fat mass. Serum was tested for alanine aminotransferase (ALT, IU/L), aspartate aminotransferase (AST, IU/L), gamma-glutamyltransferase (GGT, IU/L), high-sensitivity C-reactive protein (mg/L), hepatitis C virus antibody and RNA, and hepatitis B virus core antibody and surface antigen as described.^32^ Hepatitis C virus infection was indicated by a positive RNA or confirmed antibody test and hepatitis B virus infection by a positive surface antigen test.

### Analysis sample

Of 5,569 NHANES 2017-2018 participants 20 years and older who were interviewed at home, 5,265 attended a mobile examination center visit. Persons ineligible for liver elastography because of pregnancy or undetermined pregnancy status or other reason, such as an insulin pump or other implantable electronic device, were excluded (n=225). Participants who did not undergo liver elastography because of refusal, insufficient time or other reason, including physical or technical limitations, or who underwent an examination, but had no complete stiffness measures were also excluded (n=170). The resulting sample for analyses of liver elastography consisted of 4,870 persons (**Figure 1**). Of these, 4,510 had complete liver elastography and 394 were considered by the National Center for Health Statistics to have a partial examination because of fasting < 3 hours (n=179), 1 to <10 valid measures obtained (n=83), or a stiffness IQR/median ≥ 30% (n=98). Compared with persons with a complete examination, those who fasted < 3 hours were younger, had a lower median CAP, and were similar on other characteristics (**Supplementary Table 1**). Participants with 1 to <10 valid measures were older and had a higher BMI. As expected, they had fewer valid measures despite more attempted measures, and had higher liver stiffness median, IQR, and IQR/median, as well as a higher median CAP. Persons with a stiffness IQR/median ≥ 30% were more likely to be non-Hispanic white and had a higher BMI, more attempted measures, and a higher liver stiffness median, IQR, and IQR/median. The higher BMI, stiffness, and CAP measures of participants in the latter two groups suggested they had potentially more severe liver injury; consequently, we chose to include them in our analysis to reflect the full range of U.S. representative population liver disease intermediate outcomes and biomarkers. Analyses that included DXA body composition measures were conducted among persons aged 20-59 years with information on one or more of the DXA measures of interest (n=2,373).

**Figure 1.**
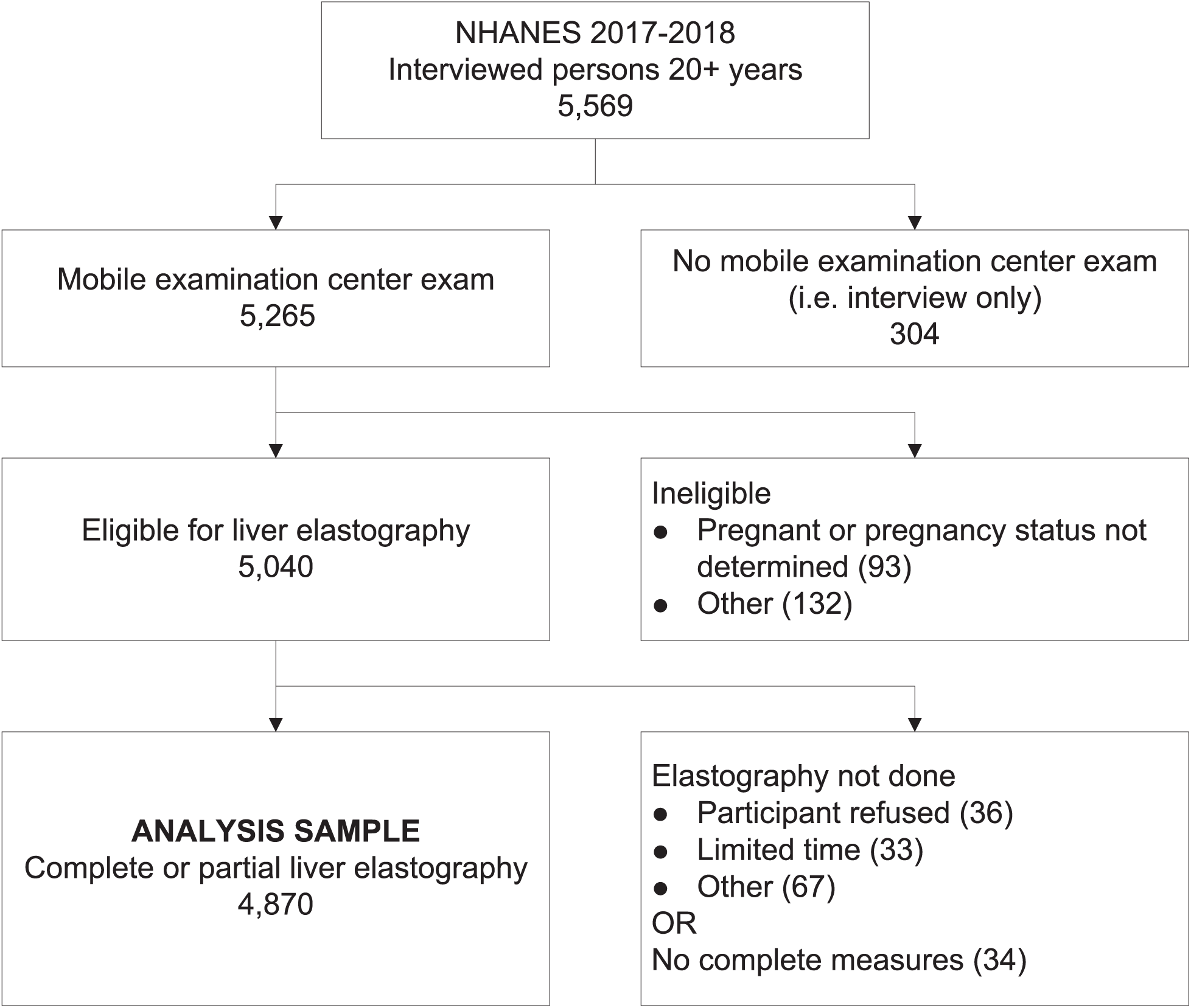
Analysis sample for NHANES 2017-2018 transient elastography NHANES, National Health and Nutrition Examination Survey.

### Statistical analysis

We first evaluated characteristics of participants with complete and partial (fasted <3 hours, 1 to <10 valid measures, or stiffness IQR/median ≥ 30%) liver elastography by comparing means (standard deviations) of continuous factors using analysis of variance and percentages of categorical factors using chi-square 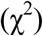 tests or linear regression. Because CAP and liver stiffness differ by sex, distributions of these measures are presented separately for men and women. Relationships of higher CAP and liver stiffness with anthropometric and DXA body composition measures were examined in unadjusted analyses using logistic regression to calculate odds ratios (OR) and 95% confidence intervals (CI) (SUDAAN PROC RLOGIST).

CAP and liver stiffness were categorized as binary measures using the 75^th^ percentiles as cut points (306.47 dB/m for CAP and 6.05 kPa for stiffness). Multiple logistic regression analysis was then used to determine the independent effects of demographics and anthropometric and DXA body composition measures on CAP and liver stiffness. Race-ethnicity, sex, and age were initially forced into models. A biological plausibility approach was followed to sequentially test anthropometric measures (BMI, BMI and waist-to-hip ratio, and waist circumference), viral hepatitis markers, liver enzymes (ALT, AST, and GGT), CAP (for models of liver stiffness), and C-reactive protein for inclusion in models.^33^ Because BMI and waist circumference were highly correlated, as were liver enzyme activities, the anthropometric measure(s) and liver enzyme that explained the highest proportion of variance (R^2^) were selected. Factors were retained in final models if p<0.10. DXA measures were evaluated in analyses including persons aged 20-59 years, the group who underwent a DXA examination.

Because there are no well-established cut points for elevated CAP and liver stiffness in the general population, we conducted sensitivity analyses in which these measures were treated as continuous characteristics using linear regression (SUDAAN PROC REGRESS). Liver stiffness was skewed to the right; therefore, natural log transformation was performed to normalize the distribution for linear regression analyses. Because they were not normally distributed, ALT, AST, GGT, CAP (for models of liver stiffness), and C-reactive protein were expressed as sex-specific deciles (10^th^ percentiles) for regression analyses using cut points shown in

**Supplementary Table 2**. Multivariate analyses excluded persons with missing values for any factor included in the model. P-values were two-sided, and P<0.05 was considered to indicate statistical significance. All analyses utilized sample weights that accounted for unequal selection probabilities and nonresponse. All variance calculations accounted for survey design effects using Taylor series linearization.^34^ SAS 9.4 (SAS Institute, Cary, NC, USA) and SUDAAN 11 (RTI, Research Triangle Park, NC, USA) were used for all analyses.

## RESULTS

Demographic characteristics, CAP and liver stiffness, liver disease history, anthropometric and DXA body composition measures, viral hepatitis markers, and liver enzyme activities for men and women are shown in **Table 1**. Compared with men, women were older, had lower CAP, liver stiffness, and liver enzyme activities, differed on all anthropometric measures, except BMI, and all DXA measures, and had higher C-reactive protein concentration and lower family income; therefore, distributions of elastography measures are presented separately for men and women. Women had a lower prevalence of health care provider diagnosed liver conditions (**Table 1**) and of fatty liver (1.9% among women vs. 2.4% among men), liver fibrosis (0.05% vs. 0.3%), liver cirrhosis (0.1% vs. 0.6%), and viral hepatitis (0.7% vs. 1.1%), but not autoimmune hepatitis (0.2% vs. 0.1%); however, the number of participants reporting diagnosed liver conditions was small and differences did not reach statistical significance.

**Table 1.** Characteristics of men and women 20+ years in the National Health and Nutrition Examination Survey, United States, 2017-2018

| Characteristic<sup>1</sup> | Men<br>(n=2,541) | Women<br>(n=2,724) | p-value |
| --- | --- | --- | --- |
| Race-ethnicity<sup>2</sup> |  |  | 0.11 |
| Non-Hispanic white | 62.1 | 62.4 |  |
| Non-Hispanic black | 10.8 | 12.1 |  |
| Non-Hispanic Asian | 5.6 | 6.2 |  |
| Hispanic | 16.2 | 15.4 |  |
| Age (years) | 47.6 (17.1) | 49.0 (17.4) | 0.004 |
| Liver elastography measures |  |  |  |
| CAP (dB/m) | 274.1 (63.4) | 254.4 (61.2) | <0.001 |
| Stiffness (kPa) | 6.4 (6.1) | 5.5 (4.6) | 0.003 |
| Liver disease history | 10.4 | 12.6 |  |
| Liver condition diagnosis ever | 5.7 | 4.2 | 0.079 |
| Still have liver condition | 3.5 | 2.4 | 0.16 |
| Duration of liver condition diagnosis (years) | 10.4 (11.5) | 12.6 (15.1) | 0.29 |
| Anthropometric measures |  |  |  |
| BMI (kg/m<sup>2</sup>) | 29.7 (6.4) | 30.0 (8.0) | 0.52 |
| Waist circumference (cm) | 103.4 (16.5) | 98.5 (17.8) | <0.001 |
| Hip circumference (cm) | 105.8 (12.2) | 109.6 (15.8) | <0.001 |
| Waist-to-hip ratio | 97.4 (7.4) | 89.6 (7.1) | <0.001 |
| DXA body composition measures<sup>3</sup> |  |  |  |
| Trunk fat mass (kg) | 12.9 (6.2) | 14.3 (7.1) | 0.036 |
| Extremity fat mass (kg)<sup>4</sup> | 11.0 (4.4) | 14.8 (5.7) | <0.001 |
| Total percent fat (%) | 27.4 (6.1) | 38.6 (6.7) | <0.001 |
| Hepatitis C virus |  |  | 0.14 |
| RNA negative, antibody positive | 0.89 | 1.09 |  |
| RNA positive | 1.68 | 0.37 |  |
| Hepatitis B virus positive | 0.30 | 0.26 | 0.68 |
| Liver enzymes (IU/L) |  |  |  |
| ALT | 27.7 (19.5) | 18.6 (12.7) | <0.001 |
| AST | 24.4 (14.5) | 20.2 (11.8) | <0.001 |
| GGT | 36.2 (45.4) | 24.3 (33.4) | <0.001 |
| C-reactive protein (mg/L) | 3.3 (7.1) | 4.5 (8.1) | <0.001 |
| Family income to poverty threshold ratio |  |  | 0.007 |
| <1.0 | 10.8 | 14.2 |  |
| 1.0-<2.0 | 19.9 | 21.6 |  |
| 2.0-<3.0 | 16.8 | 14.0 |  |
| 3.0-<5.0 | 24.0 | 24.9 |  |
| ≥5.0 | 28.5 | 25.2 |  |

CAP, controlled attenuation parameter; BMI, body mass index; DXA, dual-energy x-ray absorptiometry; RNA, ribonucleic acid; ALT, alanine aminotransferase; AST, aspartate aminotransferase; GGT, gamma-glutamyltransferase.
<sup>1</sup>Values are percentage or mean (standard deviation).
<sup>2</sup>Percentage total is less than 100% because persons of “other” race-ethnicity are not shown.
<sup>3</sup>Measured on persons 20-59 years.
<sup>4</sup>Sum of fat mass from right arm, left arm, right leg, and left leg.

### CAP

CAP ranged from 100 to 400 dB/m. Men of all racial-ethnic groups had substantially higher CAP than did women (**Table 2**). In men, CAP was significantly higher in non-Hispanic whites, non-Hispanic Asians, and Hispanics than in non-Hispanic blacks (p<0.001). In women, CAP was significantly higher in Hispanics than in non-Hispanic blacks (p<0.05). CAP generally increased with age in both men and women (**Figures 2a and 2b, Supplementary Table 3**), and markedly increased as BMI increased in both men and women (**Table 3**).

**Table 2.** Liver elastography measures by sex and race-ethnicity among participants 20+ years in the National Health and Nutrition Examination Survey, United States, 2017-2018

| Sex<br>Race | N | Mean (SD) | Geometric<br>mean ±<br>SEM<sup>1</sup> | Percentile<sup>2</sup> | | | | | | | | |
| --- | --- | --- | --- | --- | --- | --- | --- | --- | --- | --- | --- | --- |
|  |  |  |  | 5<sup>th</sup> | 10<sup>th</sup> | 15<sup>th</sup> | 25<sup>th</sup> | 50<sup>th</sup> | 75<sup>th</sup> | 85<sup>th</sup> | 90<sup>th</sup> | 95<sup>th</sup> |
| Controlled attenuation parameter (CAP; dB/m) |  |  |  |  |  |  |  |  |  |  |  |  |
| Both sexes | 4,869 | 264.1 (63.1) | 256.3 ± 1.6 | 165.1 | 186.6 | 198.2 | 216.4 | 261.1 | 306.5 | 332.7 | 351.5 | 376.7 |
| Men |  |  |  |  |  |  |  |  |  |  |  |  |
| All races<sup>3</sup> | 2,405 | 274.2 (63.4) | 266.5 ± 2.0 | 175.3 | 195.4 | 207.8 | 225.7 | 271.3 | 319.5 | 345.7 | 363.6 | 386.8 |
| NH white | 837 | 276.9 (63.1) | 269.5 ± 2.3 | 183.5 | 203.2 | 209.9 | 226.7 | 271.9 | 322.1 | 350.7 | 368.7 | 390.8 |
| NH black | 554 | 251.0 (64.5)<sup>4</sup> | 242.6 ± 4.2<sup>4</sup> | 152.4 | 171.0 | 182.6 | 202.3 | 242.5 | 293.2 | 324.6 | 347.2 | 365.1 |
| NH Asian | 339 | 275.4 (61.7) | 267.8 ± 2.6 | 180.3 | 201.6 | 213.3 | 232.1 | 273.7 | 316.3 | 346.7 | 363.2 | 378.3 |
| Hispanic | 535 | 280.5 (61.7) | 273.0 ± 3.2 | 169.5 | 192.9 | 212.2 | 239.0 | 285.3 | 320.8 | 343.8 | 359.3 | 383.4 |
| Women |  |  |  |  |  |  |  |  |  |  |  |  |
| All races<sup>3</sup> | 2,464 | 254.4 (61.2)<sup>5</sup> | 246.7 ± 2.1<sup>5</sup> | 158.2 | 178.3 | 191.3 | 210.5 | 251.2 | 294.5 | 318.9 | 336.9 | 363.8 |
| NH white | 821 | 254.0 (61.2) | 246.4 ± 3.0 | 158.6 | 177.6 | 190.7 | 210.8 | 248.8 | 293.6 | 318.7 | 337.2 | 364.6 |
| NH black | 585 | 248.8 (60.9) | 241.1 ± 2.6 | 152.9 | 173.7 | 183.3 | 204.5 | 247.5 | 288.2 | 312.0 | 329.8 | 356.4 |
| NH Asian | 363 | 250.2 (56.0) | 243.6 ± 2.8 | 157.2 | 180.5 | 194.0 | 208.4 | 247.6 | 288.6 | 311.5 | 331.2 | 343.3 |
| Hispanic | 584 | 260.9 (64.3) | 252.1 ± 4.7 | 147.8 | 182.5 | 195.8 | 218.2 | 262.1 | 304.4 | 325.3 | 340.2 | 373.1 |
| Liver stiffness (kPa) |  |  |  |  |  |  |  |  |  |  |  |  |
| Both sexes | 4,870 | 5.9 (5.4) | 5.2 ± 0.1 | 3.0 | 3.3 | 3.5 | 4.0 | 4.8 | 6.0 | 7.0 | 8.1 | 11.0 |
| Men |  |  |  |  |  |  |  |  |  |  |  |  |
| All races<sup>3</sup> | 2,406 | 6.4 (6.1) | 5.5 ± 0.1 | 3.2 | 3.6 | 3.8 | 4.2 | 5.1 | 6.3 | 7.3 | 8.7 | 12.3 |
| NH white | 837 | 6.5 (6.6) | 5.5 ± 0.1 | 3.2 | 3.6 | 3.8 | 4.2 | 5.1 | 6.3 | 7.5 | 8.8 | 13.1 |
| NH black | 555 | 6.0 (3.5) | 5.5 ± 0.1 | 3.5 | 3.7 | 4.0 | 4.3 | 5.2 | 6.4 | 7.1 | 7.8 | 10.1 |
| NH Asian | 339 | 5.9 (4.7) | 5.3 ± 0.1 | 3.2 | 3.5 | 3.7 | 4.1 | 5.0 | 6.0 | 6.7 | 7.9 | 10.4 |
| Hispanic | 535 | 6.2 (5.4) | 5.5 ± 0.1 | 3.2 | 3.6 | 3.8 | 4.3 | 5.3 | 6.2 | 7.3 | 8.6 | 10.9 |
| Women |  |  |  |  |  |  |  |  |  |  |  |  |
| All races<sup>3</sup> | 2,464 | 5.4 (4.6)<sup>6</sup> | 4.9 ± 0.1<sup>5</sup> | 2.8 | 3.1 | 3.3 | 3.7 | 4.5 | 5.8 | 6.6 | 7.6 | 9.6 |
| NH white | 821 | 5.5 (5.0) | 4.9 ± 0.1 | 2.8 | 3.1 | 3.3 | 3.7 | 4.5 | 5.8 | 6.6 | 7.7 | 9.6 |
| NH black | 585 | 5.7 (5.0) | 5.0 ± 0.1 | 2.7 | 3.1 | 3.4 | 3.8 | 4.8 | 6.0 | 7.0 | 8.0 | 10.4 |
| NH Asian | 363 | 4.8 (2.3)<sup>7</sup> | 4.5 ± 0.1<sup>7</sup> | 2.8 | 3.1 | 3.3 | 3.6 | 4.4 | 5.3 | 6.0 | 6.6 | 7.4 |
| Hispanic | 584 | 5.3 (4.0) | 4.8 ± 0.1 | 2.8 | 3.2 | 3.3 | 3.7 | 4.5 | 5.6 | 6.5 | 7.3 | 9.6 |

SD, standard deviation; SEM, standard error of mean; NH, non-Hispanic.
<sup>1</sup>Liver stiffness was not normally distributed.
<sup>2</sup>Untransformed.
<sup>3</sup>Includes persons of “other” race-ethnicity not shown.
<sup>4,7</sup>Significantly different from non-Hispanic whites: <sup>4</sup>P<0.001, <sup>7</sup>P<0.05.
<sup>5,6</sup>Significantly different from men of all race-ethnicities: <sup>5</sup>P<0.001, <sup>6</sup>P<0.05.

**Figure 2.**
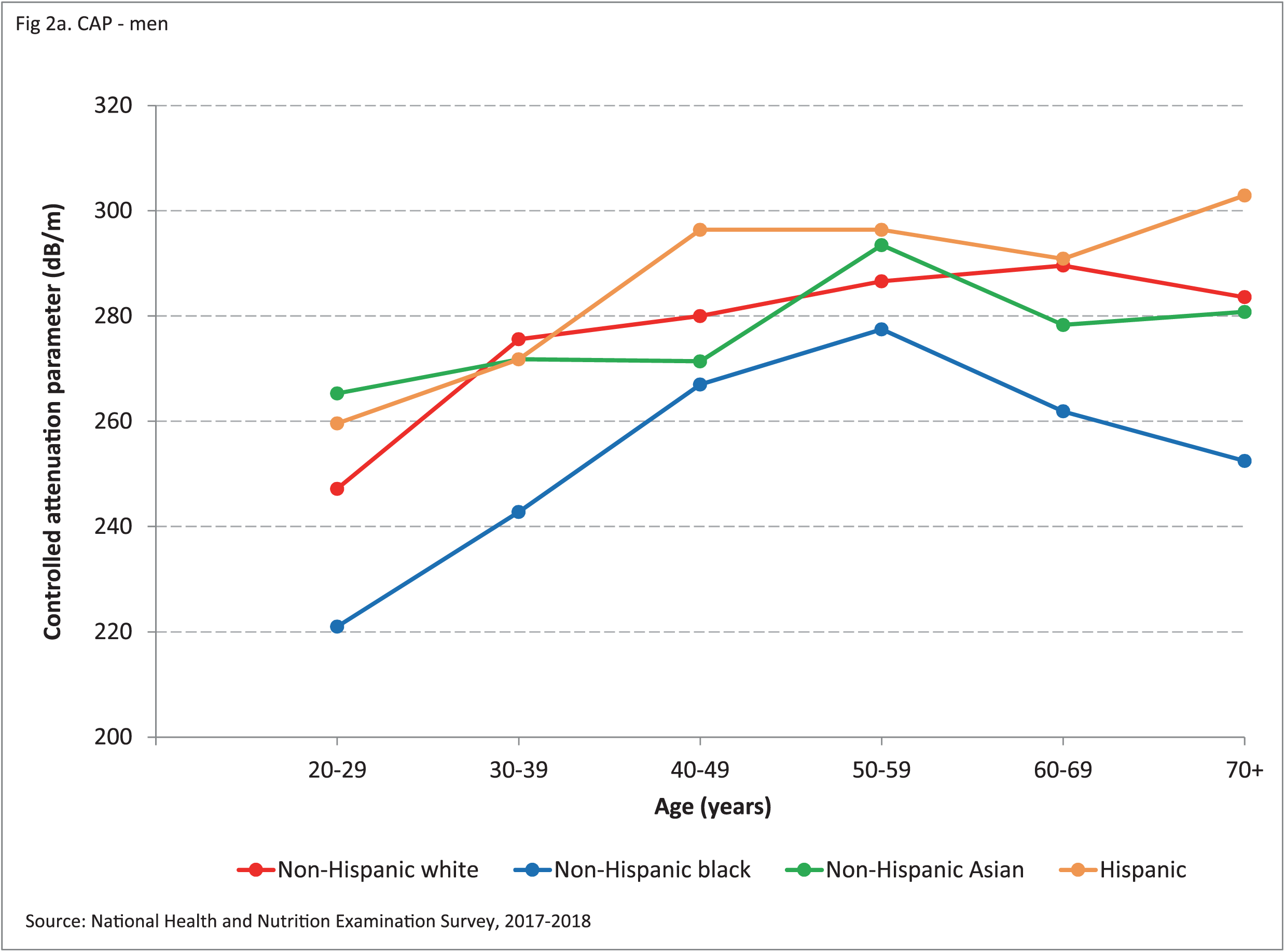
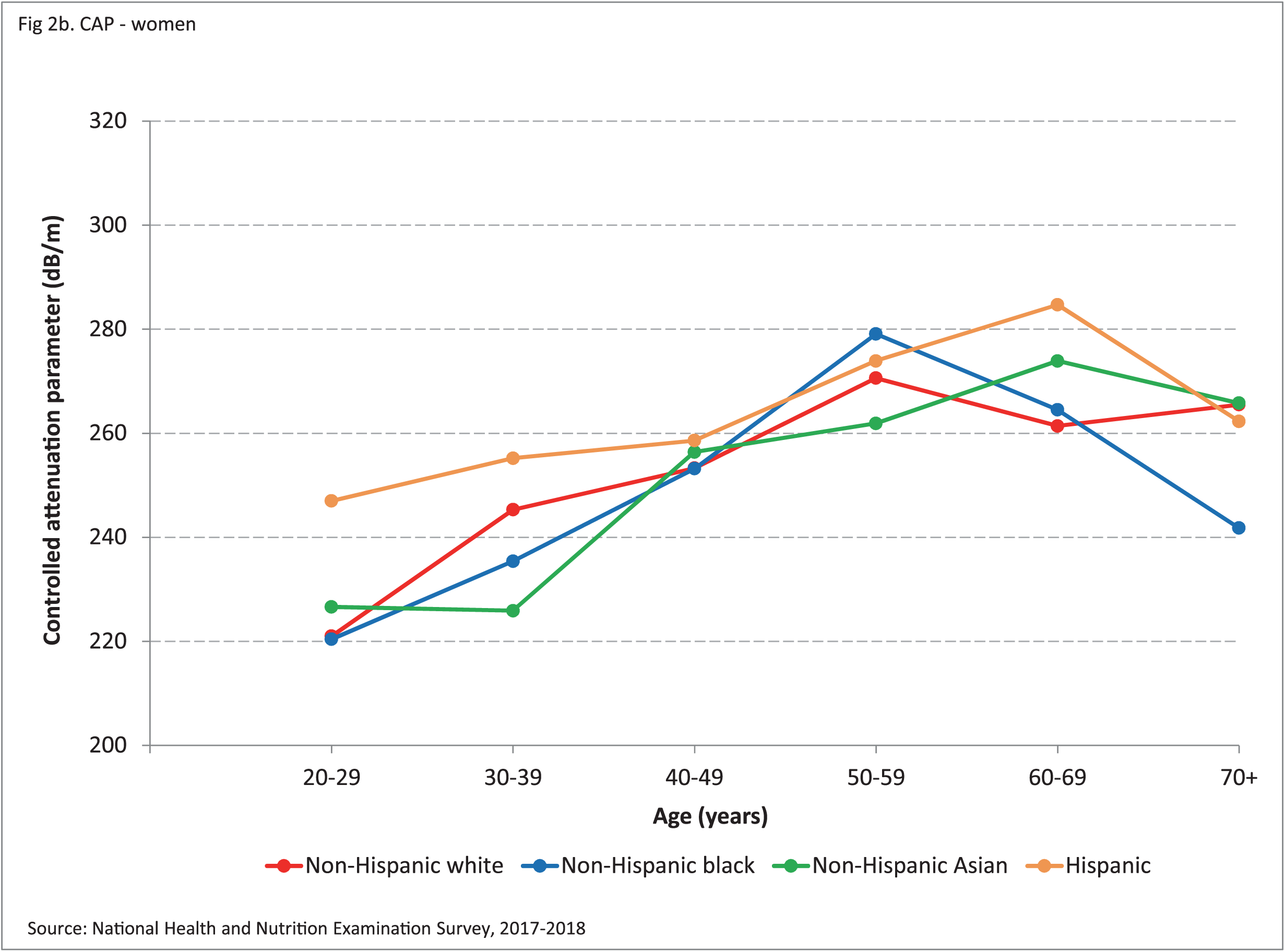
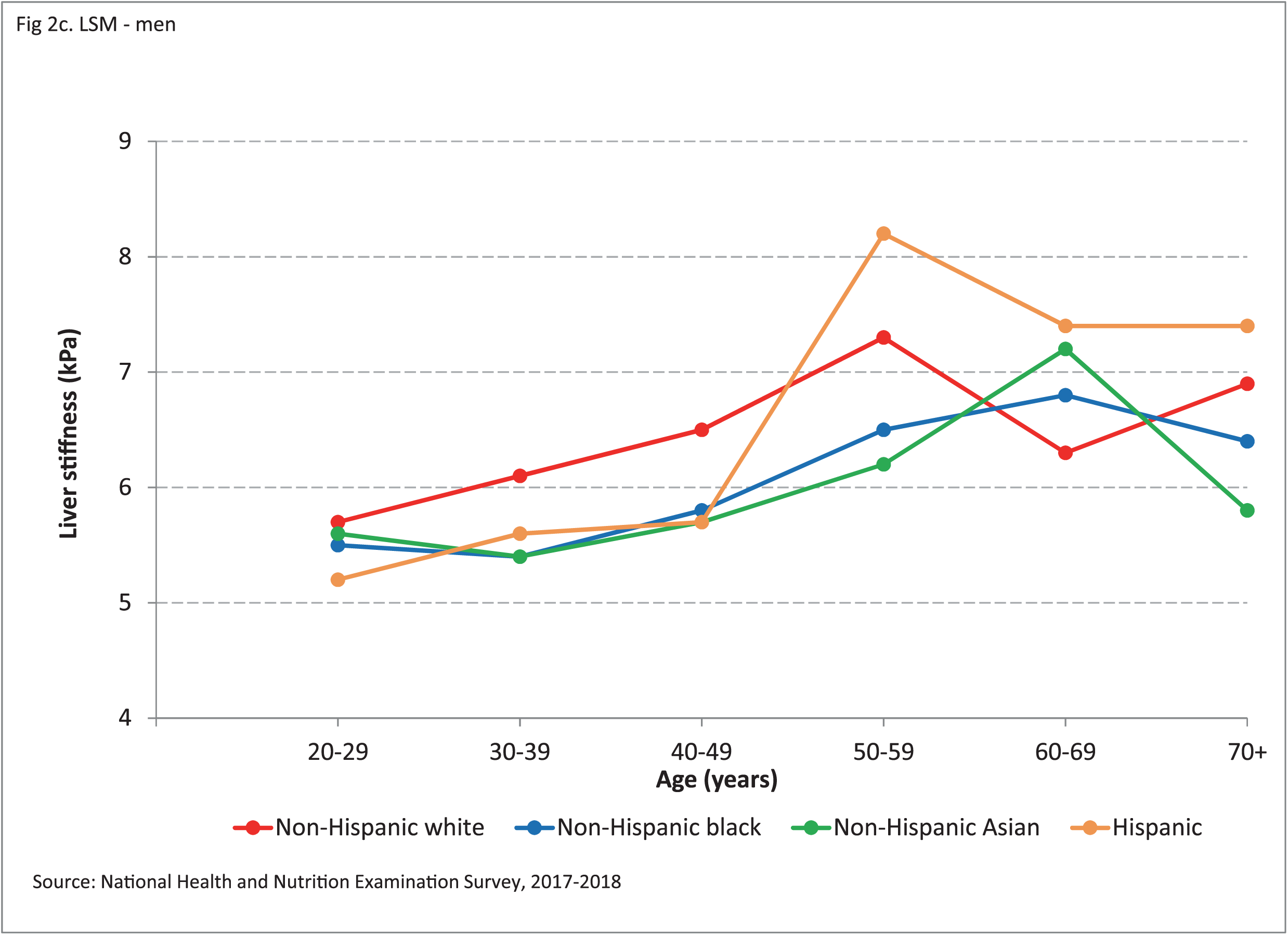
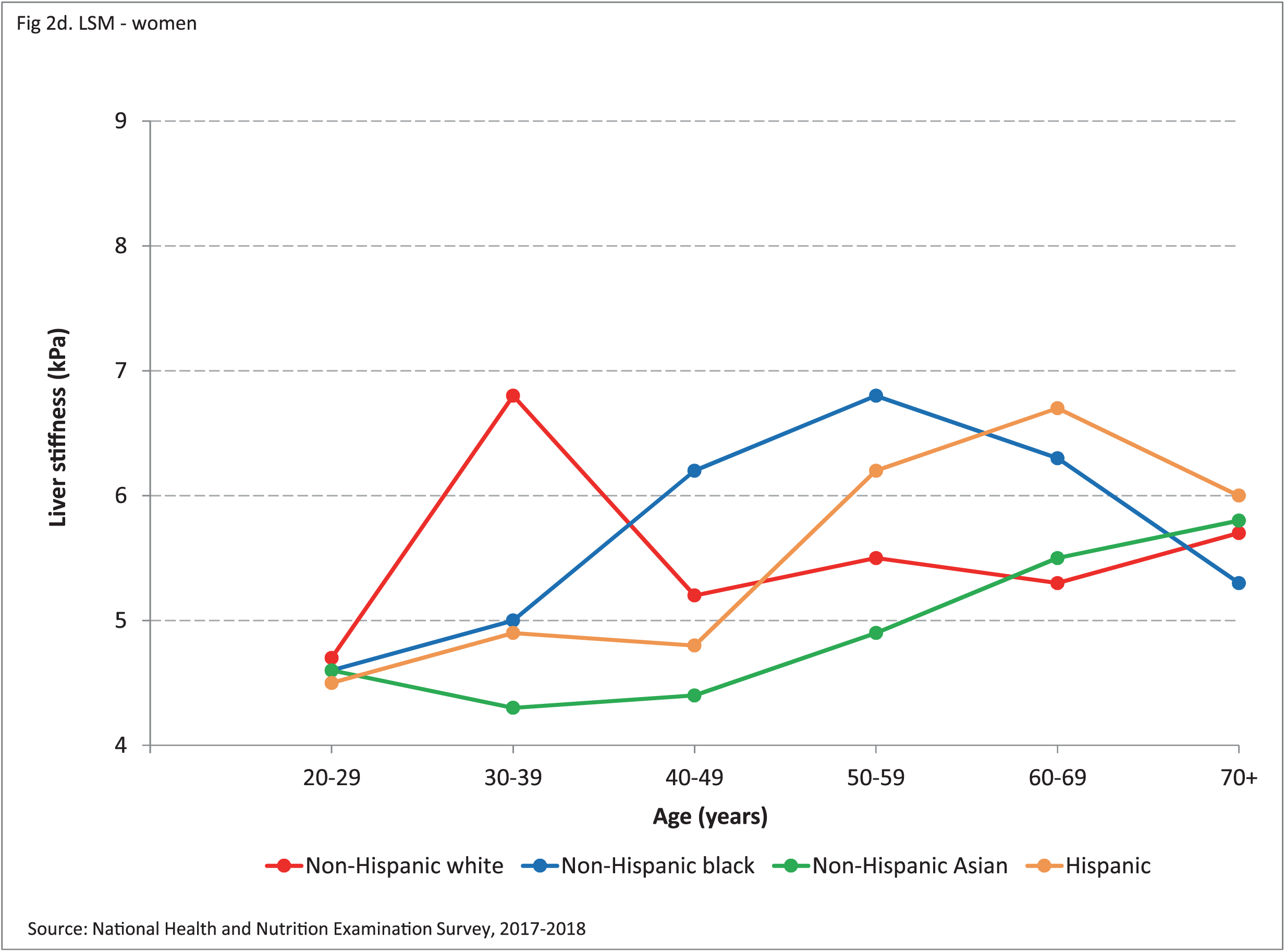
Transient elastography measures by race-ethnicity and age among participants 20+ years in the National Health and Nutrition Examination Survey, United States, 2017-2018 **(A)**. Controlled attenuation parameter among men. **(B)**. Controlled attenuation parameter among women. **(C)**. Liver stiffness among men. **(D)**. Liver stiffness among women.

**Table 3.**
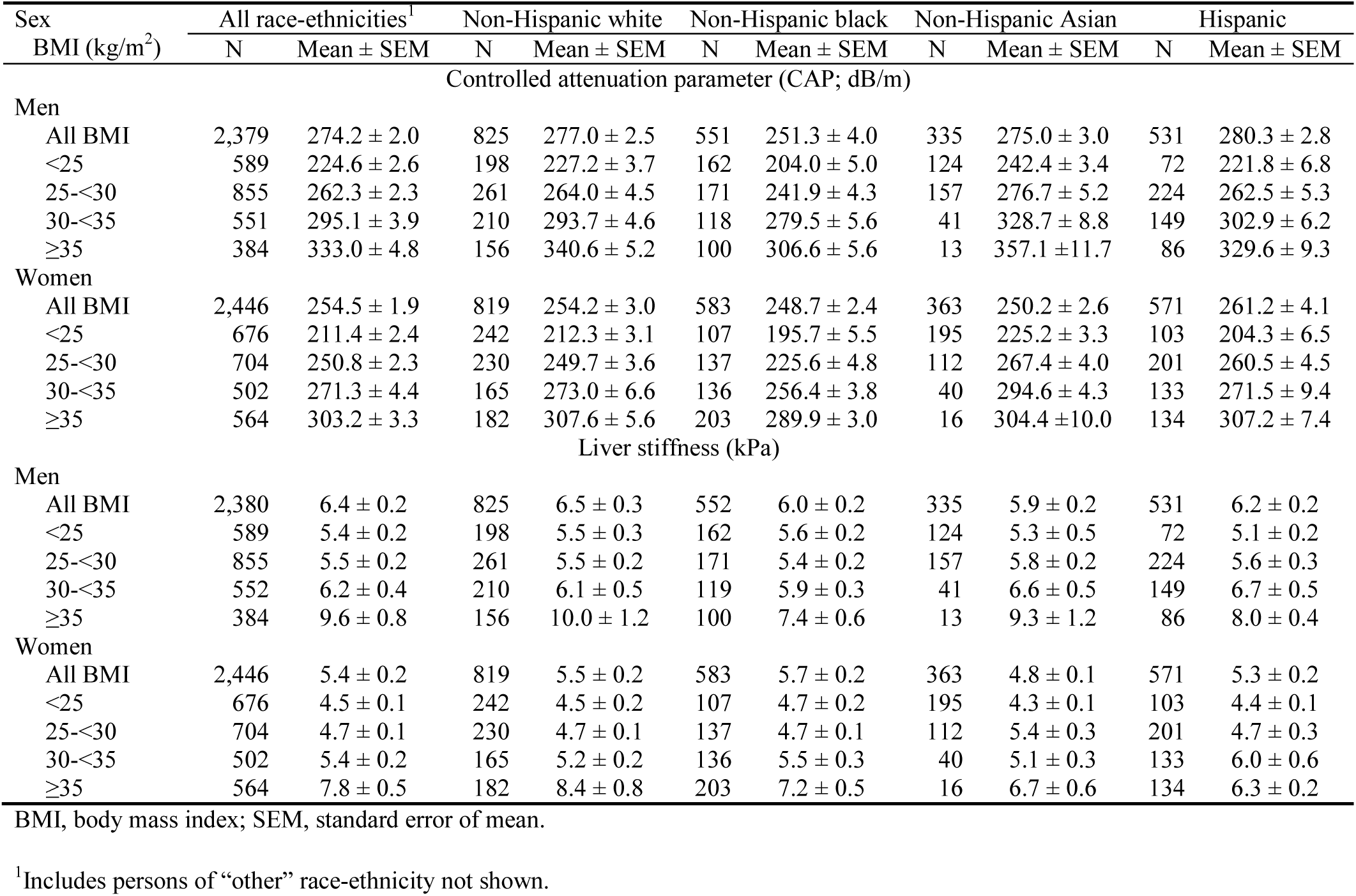
Liver elastography measures by sex, race-ethnicity, and BMI among participants 20+ years in the National Health and Nutrition Examination Survey, United States, 2017-2018

| Sex<br>BMI (kg/m<sup>2</sup>) | All race-ethnicities<sup>1</sup> |  | Non-Hispanic white |  | Non-Hispanic black |  | Non-Hispanic Asian |  | Hispanic |  |
| --- | --- | --- | --- | --- | --- | --- | --- | --- | --- | --- |
|  | N | Mean ± SEM | N | Mean ± SEM | N | Mean ± SEM | N | Mean ± SEM | N | Mean ± SEM |
| Controlled attenuation parameter (CAP; dB/m) |  |  |  |  |  |  |  |  |  |  |
| Men |  |  |  |  |  |  |  |  |  |  |
| All BMI | 2,379 | 274.2 ± 2.0 | 825 | 277.0 ± 2.5 | 551 | 251.3 ± 4.0 | 335 | 275.0 ± 3.0 | 531 | 280.3 ± 2.8 |
| <25 | 589 | 224.6 ± 2.6 | 198 | 227.2 ± 3.7 | 162 | 204.0 ± 5.0 | 124 | 242.4 ± 3.4 | 72 | 221.8 ± 6.8 |
| 25-<30 | 855 | 262.3 ± 2.3 | 261 | 264.0 ± 4.5 | 171 | 241.9 ± 4.3 | 157 | 276.7 ± 5.2 | 224 | 262.5 ± 5.3 |
| 30-<35 | 551 | 295.1 ± 3.9 | 210 | 293.7 ± 4.6 | 118 | 279.5 ± 5.6 | 41 | 328.7 ± 8.8 | 149 | 302.9 ± 6.2 |
| ≥35 | 384 | 333.0 ± 4.8 | 156 | 340.6 ± 5.2 | 100 | 306.6 ± 5.6 | 13 | 357.1 ± 11.7 | 86 | 329.6 ± 9.3 |
| Women |  |  |  |  |  |  |  |  |  |  |
| All BMI | 2,446 | 254.5 ± 1.9 | 819 | 254.2 ± 3.0 | 583 | 248.7 ± 2.4 | 363 | 250.2 ± 2.6 | 571 | 261.2 ± 4.1 |
| <25 | 676 | 211.4 ± 2.4 | 242 | 212.3 ± 3.1 | 107 | 195.7 ± 5.5 | 195 | 225.2 ± 3.3 | 103 | 204.3 ± 6.5 |
| 25-<30 | 704 | 250.8 ± 2.3 | 230 | 249.7 ± 3.6 | 137 | 225.6 ± 4.8 | 112 | 267.4 ± 4.0 | 201 | 260.5 ± 4.5 |
| 30-<35 | 502 | 271.3 ± 4.4 | 165 | 273.0 ± 6.6 | 136 | 256.4 ± 3.8 | 40 | 294.6 ± 4.3 | 133 | 271.5 ± 9.4 |
| ≥35 | 564 | 303.2 ± 3.3 | 182 | 307.6 ± 5.6 | 203 | 289.9 ± 3.0 | 16 | 304.4 ± 10.0 | 134 | 307.2 ± 7.4 |
| Liver stiffness (kPa) |  |  |  |  |  |  |  |  |  |  |
| Men |  |  |  |  |  |  |  |  |  |  |
| All BMI | 2,380 | 6.4 ± 0.2 | 825 | 6.5 ± 0.3 | 552 | 6.0 ± 0.2 | 335 | 5.9 ± 0.2 | 531 | 6.2 ± 0.2 |
| <25 | 589 | 5.4 ± 0.2 | 198 | 5.5 ± 0.3 | 162 | 5.6 ± 0.2 | 124 | 5.3 ± 0.5 | 72 | 5.1 ± 0.2 |
| 25-<30 | 855 | 5.5 ± 0.2 | 261 | 5.5 ± 0.2 | 171 | 5.4 ± 0.2 | 157 | 5.8 ± 0.2 | 224 | 5.6 ± 0.3 |
| 30-<35 | 552 | 6.2 ± 0.4 | 210 | 6.1 ± 0.5 | 119 | 5.9 ± 0.3 | 41 | 6.6 ± 0.5 | 149 | 6.7 ± 0.5 |
| ≥35 | 384 | 9.6 ± 0.8 | 156 | 10.0 ± 1.2 | 100 | 7.4 ± 0.6 | 13 | 9.3 ± 1.2 | 86 | 8.0 ± 0.4 |
| Women |  |  |  |  |  |  |  |  |  |  |
| All BMI | 2,446 | 5.4 ± 0.2 | 819 | 5.5 ± 0.2 | 583 | 5.7 ± 0.2 | 363 | 4.8 ± 0.1 | 571 | 5.3 ± 0.2 |
| <25 | 676 | 4.5 ± 0.1 | 242 | 4.5 ± 0.2 | 107 | 4.7 ± 0.2 | 195 | 4.3 ± 0.1 | 103 | 4.4 ± 0.1 |
| 25-<30 | 704 | 4.7 ± 0.1 | 230 | 4.7 ± 0.1 | 137 | 4.7 ± 0.1 | 112 | 5.4 ± 0.3 | 201 | 4.7 ± 0.3 |
| 30-<35 | 502 | 5.4 ± 0.2 | 165 | 5.2 ± 0.2 | 136 | 5.5 ± 0.3 | 40 | 5.1 ± 0.3 | 133 | 6.0 ± 0.6 |
| ≥35 | 564 | 7.8 ± 0.5 | 182 | 8.4 ± 0.8 | 203 | 7.2 ± 0.5 | 16 | 6.7 ± 0.6 | 134 | 6.3 ± 0.2 |

BMI, body mass index; SEM, standard error of mean.
<sup>1</sup>Includes persons of “other” race-ethnicity not shown.

To examine unadjusted relationships with higher CAP, the upper quartile (≥306.47 dB/m) was compared with the lower three quartiles (**Table 4**). Men were more likely to have higher CAP than women. Compared with non-Hispanic whites, non-Hispanic blacks were less likely to have higher CAP, while non-Hispanic Asians and Hispanics did not differ. Higher CAP was associated with older age. Increasing BMI, waist circumference, hip circumference, waist-to-hip ratio, trunk fat mass, extremity fat mass, total percent fat, liver enzyme activities, and C-reactive protein concentration were all associated with higher CAP. There was no relationship with hepatitis C or B virus infection or income.

**Table 4.** Unadjusted logistic regression associations of controlled attenuation parameter (CAP; dB/m) and liver stiffness (kPa) among participants 20+ years in the National Health and Nutrition Examination Survey, United States, 2017-2018

| Characteristic | CAP (n=4,869)<sup>1</sup> |  |  | Stiffness (n=4,870)<sup>2</sup> |  |  |
| --- | --- | --- | --- | --- | --- | --- |
|  | OR<sup>3</sup> | 95% CI | p-value | OR<sup>3</sup> | 95% CI | p-value |
| Sex |  |  |  |  |  |  |
| Men | 1.0 |  |  | 1.0 |  |  |
| Women | 0.56 | 0.45-0.70 | <0.001 | 0.67 | 0.56-0.79 | <0.001 |
| Race-ethnicity |  |  |  |  |  |  |
| Non-Hispanic white | 1.0 |  |  | 1.0 |  |  |
| Non-Hispanic black | 0.67 | 0.55-0.83 | 0.001 | 1.07 | 0.87-1.32 | 0.50 |
| Non-Hispanic Asian | 0.89 | 0.64-1.22 | 0.44 | 0.70 | 0.57-0.86 | 0.002 |
| Hispanic | 1.20 | 0.88-1.63 | 0.22 | 0.95 | 0.72-1.24 | 0.66 |
| Age (per 5 years) | 1.10 | 1.07-1.12 | <0.001 | 1.07 | 1.05-1.10 | <0.001 |
| Anthropometric measures |  |  |  |  |  |  |
| BMI (kg/m<sup>2</sup>) | 1.18 | 1.15-1.20 | <0.001 | 1.11 | 1.09-1.13 | <0.001 |
| Waist circumference (cm) | 1.09 | 1.08-1.10 | <0.001 | 1.05 | 1.04-1.06 | <0.001 |
| Hip circumference (cm) | 1.07 | 1.06-1.08 | <0.001 | 1.05 | 1.04-1.05 | <0.001 |
| Waist-to-hip ratio | 1.14 | 1.12-1.16 | <0.001 | 1.07 | 1.05-1.09 | <0.001 |
| DXA body composition measures<sup>4</sup> |  |  |  |  |  |  |
| Trunk fat mass (kg) | 1.21 | 1.16-1.25 | <0.001 | 1.11 | 1.08-1.14 | <0.001 |
| Extremity fat mass (kg)<sup>5</sup> | 1.15 | 1.12-1.18 | <0.001 | 1.09 | 1.06-1.11 | <0.001 |
| Total percent fat (%) | 1.07 | 1.05-1.09 | <0.001 | 1.02 | 1.01-1.04 | 0.006 |
| Hepatitis C virus |  |  |  |  |  |  |
| Negative | 1.0 |  |  | 1.0 |  |  |
| RNA-, Ab+ | 1.17 | 0.33-4.16 | 0.80 | 3.43 | 1.22-9.64 | 0.022 |
| RNA+ | 0.54 | 0.11-2.67 | 0.42 | 5.16 | 1.25-21.25 | 0.026 |
| Hepatitis B virus |  |  |  |  |  |  |
| Negative | 1.0 |  |  | 1.0 |  |  |
| Positive | 1.30 | 0.47-3.56 | 0.59 | 1.29 | 0.75-2.23 | 0.33 |
| Liver enzymes (deciles) |  |  |  |  |  |  |
| ALT | 1.27 | 1.22-1.31 | <0.001 | 1.17 | 1.12-1.22 | <0.001 |
| AST | 1.10 | 1.05-1.15 | <0.001 | 1.10 | 1.06-1.15 | <0.001 |
| GGT | 1.26 | 1.21-1.32 | <0.001 | 1.21 | 1.15-1.27 | <0.001 |
| CAP (deciles) | -- | -- | -- | 1.29 | 1.22-1.36 | <0.001 |
| C-reactive protein (deciles) | 1.28 | 1.24-1.33 | <0.001 | 1.14 | 1.10-1.19 | <0.001 |
| Family income to poverty threshold ratio |  |  |  |  |  |  |
| <1.0 | 1.17 | 0.87-1.57 | 0.27 | 1.43 | 1.06-1.93 | 0.022 |
| 1.0-<2.0 | 1.12 | 0.76-1.63 | 0.55 | 1.22 | 1.01-1.48 | 0.045 |
| 2.0-<3.0 | 1.40 | 0.91-2.13 | 0.11 | 1.65 | 1.36-2.00 | <0.001 |
| 3.0-<5.0 | 1.11 | 0.78-1.57 | 0.55 | 1.40 | 1.10-1.77 | 0.008 |
| ≥5.0 | 1.0 |  |  | 1.0 |  |  |

CAP, controlled attenuation parameter; OR, odds ratio; CI, confidence interval; BMI, body mass index; DXA, dual-energy x-ray absorptiometry; RNA, ribonucleic acid; Ab, antibody; ALT, alanine aminotransferase; AST, aspartate aminotransferase; GGT, gamma-glutamyltransferase.
<sup>1</sup>Controlled attenuation parameter was categorized as the 4<sup>th</sup> quartile ( $\geq 306.47$ dB/m) compared with the first through third quartiles.
<sup>2</sup>Liver stiffness was categorized as the 4<sup>th</sup> quartile ( $\geq 6.05$ kPa) compared with the first through third quartiles.
<sup>3</sup>From logistic regression analysis.
<sup>4</sup>Measured on persons 20-59 years.
<sup>5</sup>Sum of fat mass from right arm, left arm, right leg, and left leg.

In multivariate-adjusted analysis, men remained more likely to have higher CAP than women, although the strength of the relationship decreased slightly (**Table 5**). Compared with non-Hispanic whites, non-Hispanic blacks remained less likely to have higher CAP and Hispanics did not differ. An over two-fold greater odds of higher CAP emerged among non-Hispanic Asians with adjustment for BMI. Increasing age, BMI, waist-to-hip ratio, and ALT and C-reactive protein concentrations all remained associated with higher CAP after adjustment for all other factors in the model. When waist circumference was substituted for BMI and waist-to-hip ratio in the multivariate-adjusted model, it was associated with higher CAP (OR, 1.08; 95% CI, 1.07-1.09; p<0.001) and an association of Hispanic ethnicity with higher CAP emerged (OR, 1.65; 95% CI, 1.07-2.54; p=0.027), while other relationships remained similar. Among participants aged 20-59 years with DXA and elastography measurements, associations with higher CAP were found with substitution for BMI and waist-to-hip ratio of trunk fat mass (OR, 1.21; 95% CI, 1.15-1.28; p<0.001), extremity fat mass (OR, 1.19; 95% CI, 1.15-1.23; p<0.001), or total percent fat (OR, 1.17; 95% CI, 1.12-1.22; p<0.001) individually in the multivariate-adjusted model.

**Table 5.** Multivariate-adjusted logistic regression associations of controlled attenuation parameter (CAP; dB/m) and liver stiffness (kPa) among participants 20+ years in the National Health and Nutrition Examination Survey, United States, 2017-2018

| Characteristic | CAP (n=4,395)<sup>1</sup> |  |  | Stiffness (n=4,476)<sup>2</sup> |  |  |
| --- | --- | --- | --- | --- | --- | --- |
|  | OR<sup>3</sup> | 95% CI | p-value | OR<sup>3</sup> | 95% CI | p-value |
| Sex |  |  |  |  |  |  |
| Men | 1.0 |  |  | 1.0 |  |  |
| Women | 0.69 | 0.46-1.03 | 0.065 | 0.63 | 0.49-0.80 | <0.001 |
| Race-ethnicity |  |  |  |  |  |  |
| Non-Hispanic white | 1.0 |  |  | -- | -- | -- |
| Non-Hispanic black | 0.67 | 0.51-0.88 | 0.007 | -- | -- | -- |
| Non-Hispanic Asian | 2.32 | 1.54-3.50 | <0.001 | -- | -- | -- |
| Hispanic | 1.33 | 0.88-2.02 | 0.17 | -- | -- | -- |
| Age (per 5 years) | 1.12 | 1.08-1.17 | <0.001 | 1.07 | 1.03-1.10 | <0.001 |
| BMI (kg/m<sup>2</sup>) | 1.15 | 1.13-1.18 | <0.001 | 1.09 | 1.07-1.11 | <0.001 |
| Waist-to-hip ratio | 1.08 | 1.05-1.11 | <0.001 | -- | -- | -- |
| Hepatitis C virus |  |  |  |  |  |  |
| Negative | -- | -- | -- | 1.0 |  |  |
| RNA-, Ab+ | -- | -- | -- | 3.11 | 1.36-7.14 | 0.011 |
| RNA+ | -- | -- | -- | 6.07 | 1.14-32.35 | 0.036 |
| ALT (deciles) | 1.22 | 1.16-1.28 | <0.001 | -- | -- | -- |
| GGT (deciles) | -- | -- | -- | 1.11 | 1.05-1.16 | <0.001 |
| C-reactive protein (deciles) | 1.11 | 1.06-1.16 | <0.001 | -- | -- | -- |
| CAP (deciles) | -- | -- | -- | 1.12 | 1.07-1.18 | <0.001 |
|  | Model R<sup>2</sup>=0.30 |  |  | Model R<sup>2</sup>=0.16 |  |  |

CAP, controlled attenuation parameter; OR, odds ratio; CI, confidence interval; BMI, body mass index; RNA, ribonucleic acid; Ab, antibody; ALT, alanine aminotransferase; GGT, gamma-glutamyltransferase.
<sup>1</sup>Controlled attenuation parameter was categorized as the 4<sup>th</sup> quartile ( $\geq 306.47$ dB/m) compared with the first through third quartiles.
<sup>2</sup>Liver stiffness was categorized as the 4<sup>th</sup> quartile ( $\geq 6.05$ kPa) compared with the first through third quartiles.
<sup>3</sup>From logistic regression analysis.

Because there are no well-established cut points for elevated CAP in the general population, we conducted additional analyses with CAP treated as a continuous characteristic (**Supplementary Tables 4 and 5**). The factors associated with higher CAP in unadjusted and multivariate-adjusted models were the same as those associated in the main analyses (**Tables 4 and 5**). We also conducted sensitivity analyses limited to participants considered to have complete liver elastography (fasted ≥3 hours, ≥10 valid measures, and stiffness IQR/median <30%) (**Supplementary Tables 6 and 7**). Multivariate-adjusted results were similar to those of analyses that included participants considered to have partial elastography (**Table 5 and Supplementary Table 5**).

### Liver stiffness

Liver stiffness ranged from 1.6 to 75.0 kPa. Men of all racial-ethnic groups had substantially higher liver stiffness than did women (**Table 2**). In men, stiffness did not differ by race-ethnicity. In women, stiffness was significantly higher in non-Hispanic whites and non-Hispanic blacks than in non-Hispanic Asians (p<0.05). Liver stiffness generally increased with age in both men and women (**Figures 2 c and 2d, Supplementary Table 3**) and increased as BMI increased, especially at BMIs above 30 kg/m^2^ in both men and women (**Table 3**).

Unadjusted relationships with higher liver stiffness were examined by comparing the upper quartile (≥6.05 kPa) with the lower three quartiles (**Table 4**). Men were more likely to have higher stiffness than women. Compared with non-Hispanic whites, non-Hispanic Asians were less likely to have higher stiffness, while non-Hispanic blacks and Hispanics did not differ. Higher stiffness was associated with older age. Increasing BMI, waist circumference, hip circumference, waist-to-hip ratio, trunk fat mass, extremity fat mass, total percent fat, CAP, and liver enzyme and C-reactive protein concentrations were all associated with higher stiffness. Compared with a family income to poverty threshold ratio of ≥ 5, lower incomes were associated with higher stiffness. Though numbers of cases were small, higher stiffness was associated with both hepatitis C virus past (n=49) and current (n=51) infection. There was no relationship with hepatitis B virus infection (n=31).

In multivariate-adjusted analysis, men remained more likely to have higher stiffness than women (**Table 5**). Race-ethnicity was unrelated to higher stiffness. The decreased unadjusted odds of higher stiffness among non-Hispanic Asians was eliminated with adjustment for BMI.

Increasing age, BMI, GGT, and CAP, and hepatitis C virus infection all remained associated with higher stiffness after adjustment for all other factors in the model, while C-reactive protein was not related. Waist-to-hip ratio was not associated with higher stiffness with adjustment for BMI. When waist circumference was substituted for BMI in the multivariate-adjusted model, it was associated with higher stiffness (OR, 1.04; 95% CI, 1.03-1.05; p<0.001) and other relationships remained similar. When income was added to the multivariate model, lower incomes (except for 1.0-<2.0) were associated with higher stiffness compared with a family income to poverty threshold ratio of ≥ 5 (data not shown). Among participants aged 20-59 years with DXA and elastography measurements, associations with higher stiffness were found with substitution for BMI of trunk fat mass (OR, 1.09; 95% CI, 1.06-1.13; p<0.001) or extremity fat mass (OR, 1.08; 95% CI, 1.06-1.11; p<0.001), but not total percent fat (p=0.11) individually in the multivariate-adjusted model.

Because there are no well-established cut points for elevated liver stiffness in the general population, we conducted additional analyses with stiffness treated as a continuous characteristic (**Supplementary Tables 4 and 5**). The factors associated with higher stiffness in unadjusted and multivariate-adjusted models were the same as those associated in the main analyses (**Tables 4 and 5**) with the exception that hepatitis B virus infection was associated with higher stiffness in both unadjusted and multivariate-adjusted analyses and only the relationship with current hepatitis C virus infection was statistically significant. In sensitivity analyses limited to participants considered to have complete liver elastography (**Supplementary Tables 6 and 7**), multivariate-adjusted results were similar to those of analyses that included participants with partial elastography (**Table 5 and Supplementary Table 5**).

## DISCUSSION

In this representative sample of the U.S. adult population with transient elastography measures, we estimated distributions of CAP and liver stiffness and explored relationships with body composition. Men had higher mean values than women for both CAP and liver stiffness, and both measures increased with age and BMI in men and women. Higher CAP was strongly and positively associated with all anthropometric and DXA measures studied in unadjusted analyses and after adjustment for demographics and liver enzyme concentrations. Higher stiffness was strongly and positively associated with all anthropometric and DXA measures studied in unadjusted analyses and most associations remained with adjustment for demographics and liver enzyme concentrations. Hepatitis C virus infection was associated with higher liver stiffness, but not with higher CAP. Hepatitis B virus infection was associated with higher liver stiffness in models where liver stiffness was treated as a continuous factor.

Previous reports of CAP and liver stiffness distributions and relationships with anthropometric measures in the general population were limited to community-based samples or persons undergoing health examinations.^12,15,35–41^ Most of these studies were conducted in Europe or Asia on single ethnic groups and had smaller samples than NHANES 2017-2018. Consistent with our findings, these studies reported associations of traditional fatty liver disease risk factors, including older age, obesity, and elevated aminotransferases with increased CAP^15, 37, 38^ and liver stiffness.^36,40^ Moreover, the aging liver experiences anatomical and physiological changes that increase susceptibility to liver injury and fibrosis, including an altered inflammatory response.^42^ The progression of fatty liver disease throughout the human life span coupled with the U.S. obesity epidemic presents a significant public health threat to the health care system.

We found that non-Hispanic blacks had lower mean CAP compared with non-Hispanic whites, and non-Hispanic Asians had over twice the odds of higher CAP after adjustment for BMI. Non-Hispanic Asians had lower mean stiffness, but this relationship was eliminated with adjustment for BMI. A recent large systematic review and meta-analysis of racial and ethnic disparities in nonalcoholic fatty liver disease (NAFLD) in the United States found that the burden was highest in Hispanics and lowest in blacks.^43^ In the current analysis of NHANES 2017-2018 data, Hispanics had higher CAP compared with non-Hispanic whites without statistical significance. This slight difference may be due to our inclusion of persons with viral hepatitis or significant alcohol consumption in this initial look at transient elastography assessed liver injury in the United States or may be simply due to sample size. Data on non-Hispanic Asians in the United States are less readily available although Asians are found to develop NAFLD at lower BMI and hence may greatly benefit from further study with a larger sample size.^44^ Racial-ethnic differences observed by transient elastography merit continuation of NHANES measures to further elucidate U.S. liver health disparities and their determinants.

In NHANES 2017-2018, participants aged 20-59 years underwent whole body DXA scans. We found strong positive associations of trunk fat mass and extremity fat mass with both higher CAP and higher liver stiffness, and of total percent fat with higher CAP. Few population studies have investigated relationships of fatty liver disease with DXA body composition measures. In the Dutch Rotterdam Study, high fat mass was associated with NAFLD, with fat distribution the strongest predictor, and low skeletal muscle mass was associated with normal-weight NAFLD among women.^45^ In the Framingham Heart Study, greater central body fat was associated with increased hepatic steatosis, while greater lower extremity body fat, and in men, greater extremity lean mass were associated with decreased hepatic steatosis.^41^ Including transient elastography and DXA measures in subsequent NHANES cycles is crucial for sufficient sample sizes to study fatty liver disease and body composition within sex, race-ethnicity, and BMI groups.

Assessment of liver fibrosis in the general population has been limited to the use of prediction equations composed of biochemical and clinical measures. Transient elastography provides another non-invasive means to measure fibrosis along with steatosis. In the United States, the diagnostic accuracy of transient elastography was evaluated using liver histology as the reference standard in the multicenter NASH Clinical Research Network.^18^ Liver stiffness identified advanced fibrosis and cirrhosis with an area under the receiver operating characteristics curve of 0.83 (95% CI, 0.79-0.87) and 0.93 (95% CI, 0.90-0.97), respectively. CAP detected steatosis with an area under the curve of 0.76 (95% CI, 0.64-0.87). Transient elastography was less accurate in distinguishing lower fibrosis stages and higher steatosis grades. The investigators also found transient elastography to have low failure (3.2%) and high reliability (>95%) rates and high reproducibility in their NAFLD participants.^46^

As with previous reports that used NHANES to study noninvasive liver disease markers, it is not possible to make direct comparisons of transient elastography biomarkers with histological diagnoses that cannot be obtained in a healthy general population due to the invasive nature of liver biopsy. Thus, we were unable to evaluate cut points for clinically significant hepatic steatosis and fibrosis compared to the reference standard. Although CAP and liver stiffness cannot be validated in a general population using histology, their strong associations with obesity-related biomarkers in the current analysis provide support for their use as surrogates for liver steatosis and fibrosis. The NHANES 2017-2018 CAP and liver stiffness population distributions can be evaluated in relation to cut points identified in a multicenter study in the United Kingdom of persons with suspected NAFLD who underwent biopsy.^13^ The U.S. CAP 75^th^ percentile of 306 dB/m that we used to examine associations with higher CAP was similar to the U.K. cut point of 302 dB/m that yielded the most accurate overall prediction (Youden index) of steatosis grade greater than or equal to 1 (≥5% steatosis) (sensitivity, 0.80; specificity, 0.83). The same U.K. cut point of 302 dB/m predicted steatosis grade equal to 3 (≥67% steatosis) with a sensitivity of 0.90 (specificity, 0.38). The U.S. liver stiffness 75^th^ percentile of 6.0 kPa that we used to examine association with higher stiffness was similar to the U.K. cut point of 6.1 kPa that predicted fibrosis stage greater than or equal to 2 with a sensitivity of 0.90 (specificity, 0.38). Fibrosis stages greater than or equal to 3 and equal to 4 were predicted with a sensitivity of 0.90 by U.K. cut points of 7.1 kPa and 10.9 kPa, respectively, which were similar to the U.S. population 85^th^ and 95^th^ percentiles, respectively (Table 2).

In the current analysis, the precision of estimates, especially for population groups, is limited by the sample size using only two years of NHANES transient elastography data. Despite oversampling of racial-ethnic minorities and older adults, sample sizes for some groups were relatively small. For example, the proportion of persons aged 70 years or older was lower among minorities compared with non-Hispanic whites. Body composition changes with aging; therefore, additional survey cycles are needed to investigate body composition in older adults with elastography measures.^47^ Another limitation is the lack of data on alcohol intake. Although NHANES 2017-2018 participants were asked about alcohol intake, these data have not yet been released by the National Center for Health Statistics. Therefore, we were unable to evaluate the relationship of alcohol intake with CAP and liver stiffness. Finally, the cross-sectional design of the survey does not allow determination of causation. Despite these limitations, NHANES 2017-2018 provides the first U.S. nationally representative transient elastography biomarker data. Strengths include avoidance of ascertainment bias found in clinical studies of conveniently selected patients and ability to generalize results to U.S. population ethnic-racial groups that are differentially represented in most studies. Other benefits include the potential for larger sample size with additional NHANES survey cycles to estimate sex-, age-, ethnicity- and race-specific transient elastography biomarker cut points due to the U.S. representative multi-ethnic NHANES population and the evaluation of body composition using anthropometry and DXA measures.

In conclusion, in this report we have provided the first estimates of liver fat and fibrosis distributions assessed by transient elastography CAP and liver stiffness in the U.S. population. We also demonstrated strong positive associations of anthropometric and DXA body composition measures with higher CAP and liver stiffness. Additional cycles of NHANES transient elastography biomarker data will enable more precise estimates of distributions and relationships for increased understanding and continued monitoring of the NAFLD epidemic in the United States. Prevalence and severity of transient elastography identified NAFLD were associated with *PNPLA3* and *TM6SF2* genetic variants in a general population,^38^ and genetic polymorphisms in combination with elastography measures have been proposed for diagnosis and staging of NAFLD.^48^ Future NHANES cycles will enable exploration of genetic variants in relation to elastography steatosis and fibrosis measures in the U.S. population.

## Data Availability

All data referred to in the manuscript are publicly available.

https://www.cdc.gov/nchs/nhanes/index.htm

## ACKNOWLEDGMENTS

The authors thank Jan Walters and Lauren Prelewicz for assistance with creation of figures.

Author names in bold designate shared co-first authorship.

